# The relationship between demographic, psychosocial and health-related parameters and the impact of COVID-19: a study of twenty-four Indian regions

**DOI:** 10.1101/2020.07.27.20163287

**Authors:** Ravi Philip Rajkumar

## Abstract

**Objectives:** The impact of the COVID-19 pandemic has varied widely across nations and even in different regions of the same nation. Some of this variability may be due to the interplay of pre-existing demographic, psychological, social and health-related factors in a given population.

**Methods:** Data on the COVID-19 prevalence, crude mortality and case fatality rates were obtained from official government statistics for 24 regions of India. The relationship between these parameters and demographic, social, psychological and health-related indices in these states was examined using both bivariate and multivariate analyses.

**Results:** A variety of factors - state population, sex ratio, and burden of diarrhoeal disease and ischemic heart disease - were associated with measures of the impact of COVID-19 on bivariate analyses. On multivariate analyses, prevalence and crude mortality rate were both significantly and negatively associated with the sex ratio.

**Conclusions:** These results suggest that the transmission and impact of COVID-19 in a given population may be influenced by a number of variables, with demographic factors showing the most consistent association.

**1. What is the current understanding of this subject?**
Wide variations in the impact of COVID-19, both in terms of transmission and fatality, have been observed across nations and regions. Demographic, social and health-related factors have been implicated in this variability.
**2. What does this report add to the literature?**
This study suggests that factors such as population, sex ratio, and prior burden of certain communicable and non-communicable diseases may influence this variability within a given country.
**3. What are the implications for public health practice?**
These findings may shed light on integrated biological and psychosocial approaches to minimize the spread and mortality due to the COVID-19 pandemic.

## Introduction

The global pandemic of acute respiratory illness caused by the novel betacoronavirus SARS-CoV-2, officially designated COVID-19, has emerged as perhaps the most significant health crisis of our times.^1^ An unexpected observation in the context of this pandemic has been the existence of wide variations in the prevalence, mortality and case fatality rates across affected countries,^2,3,4^ which cannot be wholly explained on the basis of differences in the virulence of SARS-CoV-2 strains.^2,5^ While some of this variation may reflect variations in health care and testing capacity across nations,^6^ it remains important to examine the role of other factors in causing this variability, particularly socioeconomic determinants of health.^7^ There is already evidence that social factors, such as perceived sociability, socioeconomic disadvantage, health literacy, trust in regulatory authorities, and the speed and stringency of measures instituted to control the spread of COVID-19, can crucially influence these variables.^1,2,7^ These social factors interact with individual psychological responses to influence behaviour either positively or negatively: for example, an adaptive (“functional”) level of fear of COVID-19 was associated with better adherence to public health safety measures in an international sample of adults, while self-reported depression had the opposite effect.^8^ Preliminary research has found that demographic and socioeconomic factors can influence variability in the spread and impact of COVID-19 not only between countries, but within a given country; in an ecological analysis of data from the United States, poverty, number of elderly people and population density were positively correlated with COVID-19 incidence and mortality rates.^9^

At the time of writing this paper, India ranked fourth among all nations in terms of the total number of confirmed cases of COVID-19, behind the United States, Brazil and Russia, with over 600,000 cases reported as of July 2, 2020.^10^ Following the initial identification of 563 positive cases, the Indian government instituted a nation-wide lockdown for a period of 21 days, which began at midnight on March 24, 2020, and was gradually relaxed over the next two months.^11^ Data from the initial phase of the lockdown suggested that this measure significantly reduced the transmission of COVID-19^12^; however, this number rapidly increased in subsequent months. This rapid increase was not uniform: across the 32 states and territories of India, certain states have reported over 1,000 cases, while others have reported far lower numbers despite their geographical proximity to these states.^13^

Besides the demographic and socioeconomic variables discussed above, an important factor that may influence such variations in the Indian context is the availability and quality of health care. Health care facilities in India are unevenly distributed, with a significant urban-rural divide,^14^ and this inequality has been further exacerbated by the COVID-19 pandemic.^15^

Keeping the above in mind, an exploratory study was conducted to examine the relationship between demographic, socioeconomic and health-related indices and measures of the spread and impact of COVID-19 across the different states of India.

## Methods

### COVID-19-related data

The current study was an exploratory, cross-sectional study based on data officially released by the Government of India. Information related to COVID-19 was obtained from the website of the Ministry of Health and Family Welfare (https://mohfw.gov.in), which provides information on the total number of cases, active cases, recovered cases and deaths for each state and territory of India and is updated every 24 hours. Data for this study was recorded from the above source on June 30, 2020. Out of the 32 states and territories, only the 24 regions which reported at least 500 cases and one or more deaths were selected, to permit a meaningful computation of COVID-19-related indices.

After obtaining information on the population of each region from the Government of India’s official census data (available at https://censusindia.gov.in/census_and_you/area_and_population.aspx), the following indices related to COVID-19 were calculated for each state:

- The prevalence rate, defined as the total number of cases (active, recovered and deceased) per 1 million population
- The crude mortality rate, defined as the total number of reported deaths due to COVID-19 per 1 million population
- The case fatality rate, defined as the ratio of deaths to all cases with outcomes (death or recovery), expressed as a percentage.

### Demographic information

Details on population per state were recorded using the census data cited above, while information on population density was obtained from the National Institution for Transforming India (NITI-Aayog), the Government of India’s official source of data on demographic and socioeconomic variables. As age and male sex have both been associated with mortality due to COVID-19,^9,16^ mean life expectancy for each state and sex ratio per state, defined as the number of women per 1000 men, were obtained from the same source.

### Socioeconomic variables

Information on literacy rates and female literacy rates per state was obtained from official census data, while information on poverty, defined as the percentage of people living below the poverty line in each state, was obtained from the data published by the Ministry of Social Justice and Empowerment. Information on indices related to law and order – state-wise rates of homicide, accident and rape – were obtained from the official statistics published in 2018 by the National Crime Records Bureau. This information was included due to the proposed role of law enforcement, and adherence to it, in containing the spread of COVID-19.^2^

### Health-related variables

Information on a number of general indices of health for each state – the maternal, infant, and under-five mortality rates and the percentage of children under 24 months who were fully immunized – was obtained from official NITI-Aayog data, which was updated for the fiscal year 2015-16. In addition, information on the percentage of disability-adjusted life years (DALY) for six common health conditions – diarrhoeal disease, lower respiratory infection, tuberculosis, diabetes mellitus, chronic obstructive pulmonary disease, and ischemic heart disease – was obtained from the official report on state-level disease burdens commissioned by the Department of Health Research, Ministry of Health and Family Welfare and published in 2017.^17^ These variables were studied due to the emerging evidence on the role of medical comorbidities in determining the outcome of COVID-19,^18^ as well as the hypothesized role of past infectious diseases in influencing the host immune response to SARS-CoV-2.^3^ In view of the proposed relationship between depression and decreased adherence to public health measures, estimated state-wise prevalence rates of depression were obtained from the 2017 Global Burden of Disease Study.^19^

### Data analysis

Data was analyzed using the Statistical Package for Social Sciences, version 20 (SPSS 20.0, IBM Corporation). Prior to bivariate analysis, all study parameters were tested for normality. As the COVID-19 indices – prevalence, mortality and case fatality rate – were not normally distributed (p < 0.01 for all indices, Shapiro-Wilk test), Spearman’s rank correlation coefficient (ρ) was used to test the hypothesis of a monotonic relationship between these indices and the aforementioned demographic, socio-economic and health-related indices. In view of the exploratory nature of our study, a conservative significance level of p < 0.01 was used for all bivariate analyses, and all tests were two-tailed.

To confirm the strength of these associations, multivariate linear regression was carried out for each of the individual COVID-19 indices. Only those variables which were associated with these indices at least at a trend level (p < 0.05) were included in the multivariate analyses.

## Results

Data was obtained for 23 Indian states and one territory (Delhi). The mean and standard deviation values of prevalence, crude mortality rate and case fatality rate for the entire sample were 504.13 (896.64), 12.68 (30.03) and 2.77 (2.21) respectively. There was a wide range of variation in COVID-19 indices, with prevalence ranging from 61.25 (Jharkand) to 4440.03 (Delhi) per 1 million population, mortality ranging from 0.24 (Tripura) to 140.19 (Delhi) per 1 million population, and case fatality rate ranging from 0.09% (Tripura) to 7.9% (Maharashtra).

### Relationship between demographic variables and COVID-19 indices

COVID-19 prevalence for each region was significantly and negatively correlated with the sex ratio (Spearman’s ρ = −0.571, p = 0.008), and was positively associated with life expectancy, but only at a trend level (ρ = 0.469, p = 0.032). Crude mortality rate did not show any significant association with demographic variables, though there were trend-level associations with life expectancy (ρ = 0.489, p = 0.024) and sex ratio (ρ = −0.519, p = 0.017). In contrast, the case fatality rate was significantly correlated with the total population of each region (ρ = 0.526, p = 0.009).

### Relationship between socioeconomic variables and COVID-19 indices

The prevalence of COVID-19 showed a positive trend-level association with the literacy rate (ρ = 0.453, p = 0.031) and a negative trend-level association with the percentage of people living below the poverty line (ρ = −0.466, p = 0.035). No significant correlations were observed between COVID-19 mortality and case fatality rates and any socioeconomic parameter.

### Relationship between health-related variables and COVID-19 indices

COVID-19 prevalence was significantly and negatively correlated with the burden of diarrheal disease per state (ρ = −0.563, p = 0.004) and showed trend-level associations with the maternal mortality rate (ρ = −0.474, p = 0.019) and the burden of ischemic heart disease (ρ = 0.476, p = 0.019). In contrast, the mortality rate showed a significant positive correlation with the burden of ischemic heart disease (ρ = 0.535, p = 0.007) and trend-level relationships with maternal mortality rate (ρ = −0.499, p = 0.035), under-five mortality rate (ρ = −0.476, p = 0.04), and burden of diarrheal disease (ρ = −0.500, p = 0.013). None of the health-related variables were significantly associated with the case fatality rate.

### Multivariate analyses

All variables that were significantly associated with COVID-19 indices at p < 0.05 or lower were selected for multivariate linear regression analysis. For COVID-19 prevalence, only the sex ratio remained significantly and negatively associated with this parameter on multivariate analysis (t = −2.399, p = 0.037). A similar result was obtained for the COVID-19 mortality rate, with the sex ratio remaining significantly and negatively associated with this variable (t = −2.361, p = 0.04). As only a single study variable, namely the population size, was associated with the case fatality rate, multivariate analyses were not carried out in this case.

## Results

The results of this preliminary analysis found that certain demographic, socioeconomic and health-related variables were significantly related to the variability in COVID-19 prevalence, mortality and case fatality rates across 24 different regions of India. In particular, COVID-19 prevalence was associated with the sex ratio and the burden of diarrheal disease as measured by the percentage of DALYs associated with this disorder; COVID-19 mortality was associated with the burden of ischemic heart disease; and COVID-19 case fatality rate was associated with the total population of each region. On multivariate analysis, only the negative association between the sex ratio and COVID-19 prevalence and mortality remained significant.

The association between the sex ratio and measures of the impact of COVID-19 is in line with existing research findings. Several clinical case series, both from India and other countries, have reported a preponderance of male patients in hospitalized samples, as well as a link between male sex and mortality due to COVID-19.^20,21,22,23^ This phenomenon may be partly explained by sex differences in the immune and inflammatory response to SARS-CoV-2 infection^16^; however, in the Indian context, this relationship could also be influenced by traditionally-defined gender roles.^24^ These are associated with comparatively greater freedom of movement for men,^25^ which places them at a higher risk of exposure to infection.

Similarly, the link between state-wide differences in the burden of ischemic heart disease and mortality due to COVID-19 is supported by clinical research, which has found an association between the presence of ischemic heart disease and the severity of COVID-19.^26,27^ Moreover, ischemic heart disease is commonly associated with other medical conditions, such as systemic hypertension and chronic renal disease, which themselves worsen the outcome of COVID-19,^28^ and COVID-19 has been documented to trigger myocardial injury in patients with pre-existing coronary artery disease.^29^ No such significant association was found in this study for other medical comorbidities, such as diabetes mellitus (ρ = 0.244, p = 0.328) or chronic obstructive pulmonary disease (ρ = 0.262, p = 0.217), suggesting a certain degree of specificity for this association.

Though this could not be confirmed by multivariate analysis, population was positively correlated with the case fatality rate across the different regions of India. This association does not appear to be mediated solely by over-crowding, as no significant association was found between population density and case fatality (ρ = 0.370, p = 0.091). A possible explanation for this finding is the unequal distribution and accessibility of healthcare facilities in India, particularly in highly populated or rural areas;^30,31^ Such inequalities may lead to delays in obtaining appropriate treatment.^32^

The negative association found between the burden of diarrheal disease and the prevalence of COVID-19 across regions is an unexpected finding, as no such association was found for respiratory diseases such as lower respiratory infection (ρ = −0.227, p = 0.256) or pulmonary tuberculosis (ρ = 0.024, p = 0.911). While it has been postulated that prior exposure to respiratory coronaviruses may moderate the impact of SARS-CoV-2 infection,^3^ no such association has been suggested or demonstrated thus far for gastrointestinal infections. However, *in vitro* research has shown that intestinal replication may contribute to the progression of SARS-CoV-2 infection^33^; therefore, it is possible that prior intestinal viral infections, which are a common cause of diarrheal disease, could influence this process. Alternately, this association may simply represent a false-positive finding arising from the exploratory nature of the analyses conducted.

A number of other associations were observed at a trend level. While the direction of these associations was unexpected in some cases – such as a positive association between COVID-19 prevalence and literacy, and a negative association between COVID-19 and levels of poverty and maternal and under-five mortality rates – these findings must be interpreted with caution, owing to their low statistical significance and the large number of potential confounding factors, as well as the possibility of type I error.

The results of this study must be viewed in the light of certain limitations. First, data on demographic, socioeconomic and health-related was obtained from official Government statistics and populations which preceded the onset of the COVID-19 outbreak by a period of three to six years. Therefore, some of this information may not accurately reflect the contemporary situation in the different states of India. Second, this study did not take into account other factors that could influence the spread of COVID-19, such as cultural norms and practices, local variations in climate and temperature, and the efficiency of implementation of quarantine and related measures.1,2 Third, the data analysis did not take into account the confounding effects of other variables on the bivariate analyses. Fourth, due to logistic and manpower constraints on testing and case finding, the officially reported statistics on COVID-19 may underestimate the true scope of this problem in India.^6,13^ Finally, owing to the cross-sectional nature of this study, it was not possible to assess the relationship between the study variables and trends in the spread of COVID-19, such as the rate of increase in the number of cases.

## Conclusion

The results of this study, though preliminary and limited by the nature of the exploratory analyses, suggest that some of the factors that have been found to influence the outcome of COVID-19 at a clinical level, such as male sex and comorbid ischemic heart disease, also have an impact at the population level. Other unexpected findings, such as the link between population and case fatality and between diarrheal disease burden and COVID-19 prevalence, may represent potential socioeconomic or biological mechanisms that require further elucidation. Further longitudinal research with more sophisticated statistical modelling and up-to-date data may clarify the role of these and other demographic, socioeconomic and health-related variables in moderating the impact of COVID-19 within nations, and may inform future strategies to curtail the impact of this pandemic.

## Data Availability

The data that support the findings of this study are derived from the following sources available in the public domain:

1. The Government of India's official census data on population, sex ratio and literacy rates, accessible at https://censusindia.gov.in
2. The Government of India's National Institution for Transforming India (NITI-Aayog) official data on population density, life expectancy, mortality rates and immunization coverage, accessible at https://www.niti.gov.in
3. The Government of India's National Crime Records Bureau (NCRB) statistics on reported rates of homicide, accident and rape, accessible at https://ncrb.gov.in
4. The Government of India's Ministry of Social Justice and Empowerment statistics on poverty accessible at http://socialjustice.nic.in
5. The India State-Level Disease Burden Initiative data accessible at https://phfi.org

## Data availability statement

1. The Government of India’s official census data on population, sex ratio and literacy rates, accessible at https://censusindia.gov.in
2. The Government of India’s National Institution for Transforming India (NITI-Aayog) official data on population density, life expectancy, mortality rates and immunization coverage, accessible at https://www.niti.gov.in
3. The Government of India’s National Crime Records Bureau (NCRB) statistics on reported rates of homicide, accident and rape, accessible at https://ncrb.gov.in
4. The Government of India’s Ministry of Social Justice and Empowerment statistics on poverty, accessible at http://socialjustice.nic.in
5. The India State-Level Disease Burden Initiative data, accessible at https://phfi.org

